# Completion of electronic nursing documentation of inpatient admission assessment: insights from Australian metropolitan hospitals

**DOI:** 10.1101/2021.06.16.21259005

**Authors:** Danielle Ritz Shala, Aaron Jones, Greg Fairbrother, Duong Thuy Tran

## Abstract

**Introduction:** Electronic nursing documentation is an essential aspect of inpatient care and multidisciplinary communication. Analysing data in electronic medical record (eMR) systems can assist in understanding clinical workflows, improving care quality, and promoting efficiency in the healthcare system. This study aims to assess timeliness of completion of an electronic nursing admission assessment form and identify patient and facility factors associated with form completion in three metropolitan hospitals.

**Materials and Methods:** Records of 37,512 adult inpatient admissions (November 2018-November 2019) were extracted from the hospitals’ eMR system. A dichotomous variable descriptive of completion of the nursing assessment form (Yes/No) was created. Timeliness of form completion was calculated as the interval between date and time of admission and form completion. Univariate and multivariate multilevel logistic regression were used to identify factors associated with form completion.

**Results:** An admission assessment form was completed for 78.4% (n=29,421) of inpatient admissions. Of those, 78% (n=22,953) were completed within the first 24 hours of admission, 13.3% (n=3,910) between 24-72 hours from admission, and 8.7% (n=2558) beyond 72 hours from admission. Patient length of hospital stay, admission time, and admitting unit’s nursing hours per patient day were associated with form completion. Patient gender, age, and admitting unit type were not associated with form completion.

**Discussion:** Form completion rate was high, though more emphasis needs to be placed on the importance of timely completion to allow for adequate patient care planning. Staff education, qualitative understanding of delayed form completion, and streamlined guidelines on nursing admission and eMR use are recommended.

**Statement on conflicts of interest:** Declarations of interest: none

**Summary Table:** What was already known on the topic

- Electronic clinical documentation is an essential aspect of inpatient care and multidisciplinary communication
- Timely completion of documentation enables prompt team communication and facilitates quality patient care planning
- Australian hospitals are moving towards fully adopting eMR-based technologies to manage patient care processes and multidisciplinary communication

What this study added to our knowledge

- The timeliness of electronic nursing documentation can be improved
- There are patient- and facility-related factors which influence the completion of electronic nursing forms
- The application of advanced modelling techniques to existing eMR data assists in understanding clinician practices, processes and workflows

## 1. Introduction

Admission marks the start of a patient’s journey in the hospital inpatient setting. Patient assessment is a foundational nursing function, which involves the systematic and continuous collection of patient clinical and historical information, documentation of collected data and communication with team members^1^. Timely completion of necessary documents enables expedited team communication and facilitates quality patient care planning.^2^ Failure to do so can jeopardise optimal care. In Australia, the National Safety and Quality Health Service (NSQHS)^3^ Communicating for Safety Standard emphasises that timely, purpose-driven, and effective communication and documentation supports continuous, coordinated and safe care for patients.

Electronic medical record (eMR) systems have the potential to improve the quality of healthcare, streamline clinical workflows and increase efficiency in the healthcare system^4-7^. While the eMR has advantages, it also comes with disadvantages such as over-documentation, decreased speed of documentation, and the cost of implementation and maintenance^8^. The amount of time spent by nurses in electronic documentation is substantial and varies by nursing shifts, unit types, and unit activities^9-12^. A Korean study demonstrated that completion of electronic documentation happens outside of working hours when nurses are new, working on a day shift, or when task-based activities are unpredictable^11^.

There is limited research on nursing completion of electronic admission forms. Variations in healthcare form completion relating to age, sex, and clinical specialty have been reported.^13-15^ Younger patient age has been shown to be linked to lower completion.^14,15^ These studies explored factors relating to completion of a form, however the results related mostly to advanced care directives, rather than admission forms.^14-16^ Research on form completion has focused on physician practices, paper-based processes and electronic documentation of nursing tasks as a whole, as opposed to a single foundational process such as admission. Little is understood about the factors influencing adult admission documentation and there are no studies describing timeliness of electronic nursing admission documentation.

Extracting knowledge from big data and process mining has been examined and promoted in recent years to inform the execution of workflow processes, check conformance with guidelines, and find improvement opportunities in healthcare and nursing.^17-21^ This study focused on an electronic nursing admission assessment form which is one of the most common forms for communicating nursing care in Australia. This is important particularly because more Australian hospitals are moving towards fully adopting eMR-based technologies to manage patient care processes and multidisciplinary communication. The aim of this study was to explore and describe the timeliness of completing an electronic adult admission assessment form in a large Australian local health district, and identify factors associated with its completion. This provides data-driven insights into current nursing practice on electronic documentation of admission and informs future practice.

## 2. Materials and Methods

### 2.1 Design

This study was a retrospective review of Adult Admission Assessment (AAA) form completion. In our local health district, completion of this form is one of the routine nursing tasks upon admission into an inpatient hospital setting.

### 2.2 Sample and Setting

The study focused on an electronic AAA form in three metropolitan public hospitals within Sydney Local Health District (SLHD), New South Wales, Australia. SLHD encompasses all public hospitals and healthcare facilities in the central Sydney metropolitan area. From April to June 2020 alone, it had a total of 35,144 admitted patient episodes, and 81,606 acute overnight bed days^22^. Electronic patient information in these hospitals is primarily managed via SLHD’s Cerner Millennium eMR.

The study included 37,512 adult inpatient admissions to 43 clinical units from November 2018 to November 2019. ICU areas were included despite the existence of a parallel eMR system. Admissions to outpatient or community-based facilities, mental health, paediatric, emergency, and maternity settings were excluded. Ethical approval was obtained from the Sydney Local Health District Human Research Ethics Committee.

### 2.3 Data Collection

Records descriptive of patient admissions and AAA form completion were extracted from the eMR using the AAA form audit report. This report was built into the eMR by SLHD’s Nursing Informatics and eMR clinical application teams. The audit report was generated in spreadsheet format with each row representing an admission encounter. De-identified data fields in the audit report used for this study included patient-related characteristics (unique ID of patient, age, sex), admission details (unique ID of admission episode, hospital ward/clinical unit, date and time of admission, date and time of discharge), and AAA form details (unique ID of form, whether a form was completed, date and time of form completion).

### 2.4 Data Analysis

#### 2.4.1 Unit of Data Analysis

A nursing admission assessment form is completed for each episode of an inpatient admission. The unit of analysis was inpatient admission, multiple admissions from the same patient were treated independently.

#### 2.4.2 Study Outcomes

Within the eMR, the built-in AAA audit report populated affirmative status of form completion on the basis of whether an AAA form was activated and signed-off for a patient. If this was the case, date and time of form completion was then recorded. Completion timeliness was calculated as the time interval from patient admission time to form completion time.

#### 2.4.3 Patient-related characteristics

Patient-related characteristics were patient age, sex, length of stay, and time of admission (see Table 1). Length of stay was calculated as the time interval between time and date of admission to time and date of discharge. This was grouped into 10 categories representing the number of days and weeks in hospital. The time of admission was grouped according to typical nursing shifts in study hospitals, namely morning (7am to 1pm), evening (1 pm to 9pm), and night shift (9pm to 7am).

**Table 1:** Summary of inpatient admission encounters (November 2018-2019)

|  | ***Admission Encounters***<br>***(N=37512)*** |
| --- | --- |
| **Sex** |  |
| Female | 17741 (47.3%) |
| Male | 19771 (52.7%) |
| **Age (years)** |  |
| Mean (SD) | 63.9 (20.5) |
| Median | 68.0 |
| **Time of admission encounter (shift)** |  |
| Morning (7am-1pm) | 13613 (36.3%) |
| Evening (1pm-9pm) | 16073 (42.8%) |
| Night (9pm-7am) | 7826 (20.9%) |
| **Nursing Hours Per Patient Day (NHPPD)** |  |
| NHPPD 5 | 454 (1.2%) |
| NHPPD 5.5 | 5460 (14.6%) |
| NHPPD 6 | 25039 (66.7%) |
| Non-NHPPD | 2936 (7.8%) |
| Critical Care | 2015 (5.4%) |
| High Acuity | 1608 (4.3%) |

**Table 2:**
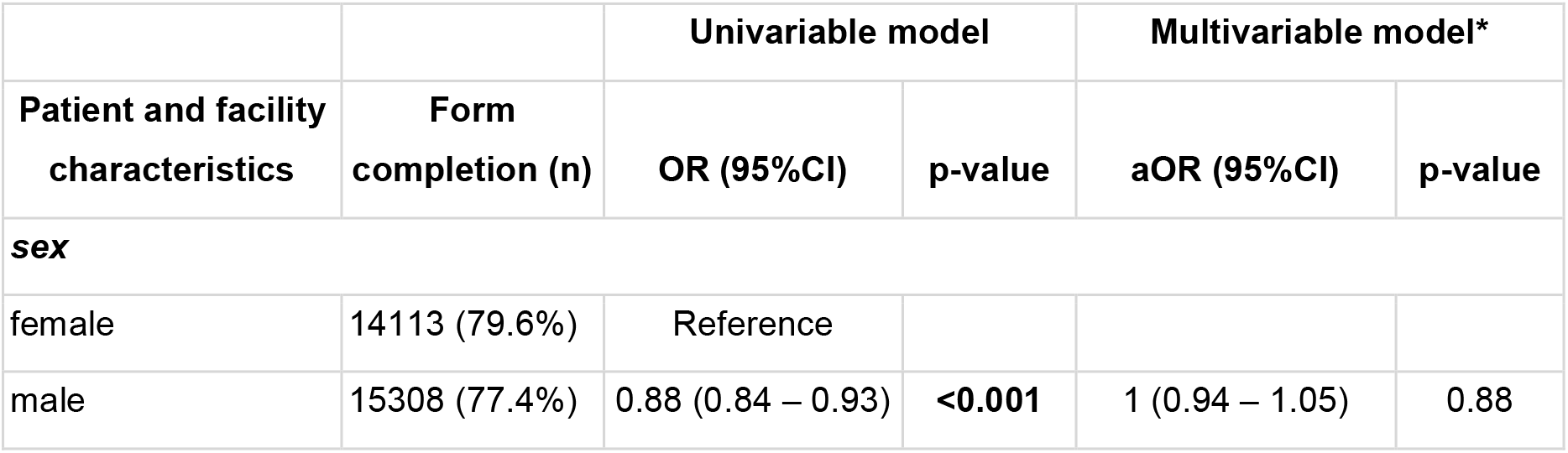
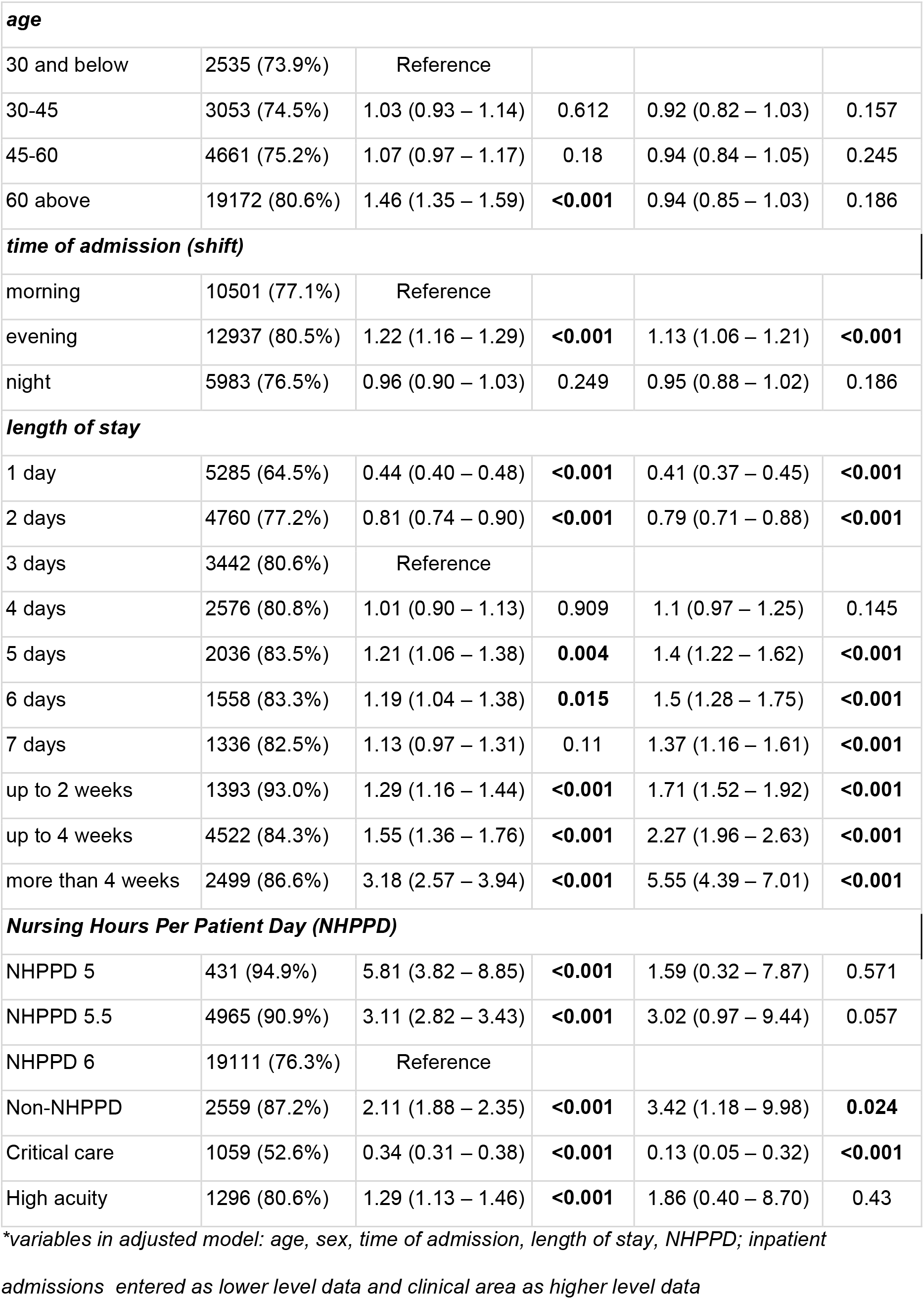
Association between form completion and patient and facility characteristics

|  |  | Univariable model |  | Multivariable model* |  |
| --- | --- | --- | --- | --- | --- |
| Patient and facility characteristics | Form completion (n) | OR (95%CI) | p-value | aOR (95%CI) | p-value |
| sex |  |  |  |  |  |
| female | 14113 (79.6%) | Reference |  |  |  |
| male | 15308 (77.4%) | 0.88 (0.84 – 0.93) | <0.001 | 1 (0.94 – 1.05) | 0.88 |
| **age** |  |  |  |  |  |
| 30 and below | 2535 (73.9%) | Reference |  |  |  |
| 30-45 | 3053 (74.5%) | 1.03 (0.93 – 1.14) | 0.612 | 0.92 (0.82 – 1.03) | 0.157 |
| 45-60 | 4661 (75.2%) | 1.07 (0.97 – 1.17) | 0.18 | 0.94 (0.84 – 1.05) | 0.245 |
| 60 above | 19172 (80.6%) | 1.46 (1.35 – 1.59) | **<0.001** | 0.94 (0.85 – 1.03) | 0.186 |
| **time of admission (shift)** |  |  |  |  |  |
| morning | 10501 (77.1%) | Reference |  |  |  |
| evening | 12937 (80.5%) | 1.22 (1.16 – 1.29) | **<0.001** | 1.13 (1.06 – 1.21) | **<0.001** |
| night | 5983 (76.5%) | 0.96 (0.90 – 1.03) | 0.249 | 0.95 (0.88 – 1.02) | 0.186 |
| **length of stay** |  |  |  |  |  |
| 1 day | 5285 (64.5%) | 0.44 (0.40 – 0.48) | **<0.001** | 0.41 (0.37 – 0.45) | **<0.001** |
| 2 days | 4760 (77.2%) | 0.81 (0.74 – 0.90) | **<0.001** | 0.79 (0.71 – 0.88) | **<0.001** |
| 3 days | 3442 (80.6%) | Reference |  |  |  |
| 4 days | 2576 (80.8%) | 1.01 (0.90 – 1.13) | 0.909 | 1.1 (0.97 – 1.25) | 0.145 |
| 5 days | 2036 (83.5%) | 1.21 (1.06 – 1.38) | **0.004** | 1.4 (1.22 – 1.62) | **<0.001** |
| 6 days | 1558 (83.3%) | 1.19 (1.04 – 1.38) | **0.015** | 1.5 (1.28 – 1.75) | **<0.001** |
| 7 days | 1336 (82.5%) | 1.13 (0.97 – 1.31) | 0.11 | 1.37 (1.16 – 1.61) | **<0.001** |
| up to 2 weeks | 1393 (93.0%) | 1.29 (1.16 – 1.44) | **<0.001** | 1.71 (1.52 – 1.92) | **<0.001** |
| up to 4 weeks | 4522 (84.3%) | 1.55 (1.36 – 1.76) | **<0.001** | 2.27 (1.96 – 2.63) | **<0.001** |
| more than 4 weeks | 2499 (86.6%) | 3.18 (2.57 – 3.94) | **<0.001** | 5.55 (4.39 – 7.01) | **<0.001** |
| **Nursing Hours Per Patient Day (NHPPD)** |  |  |  |  |  |
| NHPPD 5 | 431 (94.9%) | 5.81 (3.82 – 8.85) | **<0.001** | 1.59 (0.32 – 7.87) | 0.571 |
| NHPPD 5.5 | 4965 (90.9%) | 3.11 (2.82 – 3.43) | **<0.001** | 3.02 (0.97 – 9.44) | 0.057 |
| NHPPD 6 | 19111 (76.3%) | Reference |  |  |  |
| Non-NHPPD | 2559 (87.2%) | 2.11 (1.88 – 2.35) | **<0.001** | 3.42 (1.18 – 9.98) | **0.024** |
| Critical care | 1059 (52.6%) | 0.34 (0.31 – 0.38) | **<0.001** | 0.13 (0.05 – 0.32) | **<0.001** |
| High acuity | 1296 (80.6%) | 1.29 (1.13 – 1.46) | **<0.001** | 1.86 (0.40 – 8.70) | 0.43 |

*\*variables in adjusted model: age, sex, time of admission, length of stay, NHPPD; inpatient*
*admissions entered as lower level data and clinical area as higher level data*

#### 2.4.4 Facility-related characteristics

The facility-related characteristics were hospital, unit type, and nursing hours per patient day (NHPPD). A total of 43 clinical units were included in the study. These were classified into five unit types: medical, surgical, mixed medical-surgical, critical care, and other. Each of the clinical units in the study had a corresponding NHPPD classification supplied by the district’s nursing workforce department. The NHPPD indicator is an Australian system which monitors nursing workload within peer-to-peer public hospital groupings.^23,24^ This classification reflects the direct clinical care hours required and provided by nursing staff and takes account of diversity, complexity, and nursing tasks required within a ward. Each clinical unit in this study was assigned a NHPPD category, including NHPPD 6 (high complexity peer group A or B), NHPPD 5.5 (high complexity peer group C), and NHPPD 5 (moderate complexity). Clinical units outside of these categories fall under Critical Care, High Acuity (i.e. coronary care units), or Non-NHPPD (e.g. medical assessment, aged care, and palliative care units) classification.

#### 2.4.5 Statistical Modelling

For completion timeliness, descriptive statistics (frequency, percentage, mean, standard deviation [SD] and median) were generated. To account for within-ward variation in form completion, multilevel logistic regression was used to examine factors associated with form completion. In the multilevel modelling procedures, inpatient admission was entered as lower level data and clinical ward was entered as higher level. Both crude and adjusted odds ratio (aOR) and 95% confidence interval (CI) were estimated. Due to high correlation between unit type and NHPPD, the multivariable model included patient age, sex, length of stay, time of admission, and NHPPD. R programming language^25^ was used for data preparation and analysis. Significance tests were two-sided with alpha set at 0.05.

## 3. Results

### 3.1 Patient and facility characteristics

Patient characteristics of the sample are summarised in Table 1 and Figure 1.

**Figure 1:**
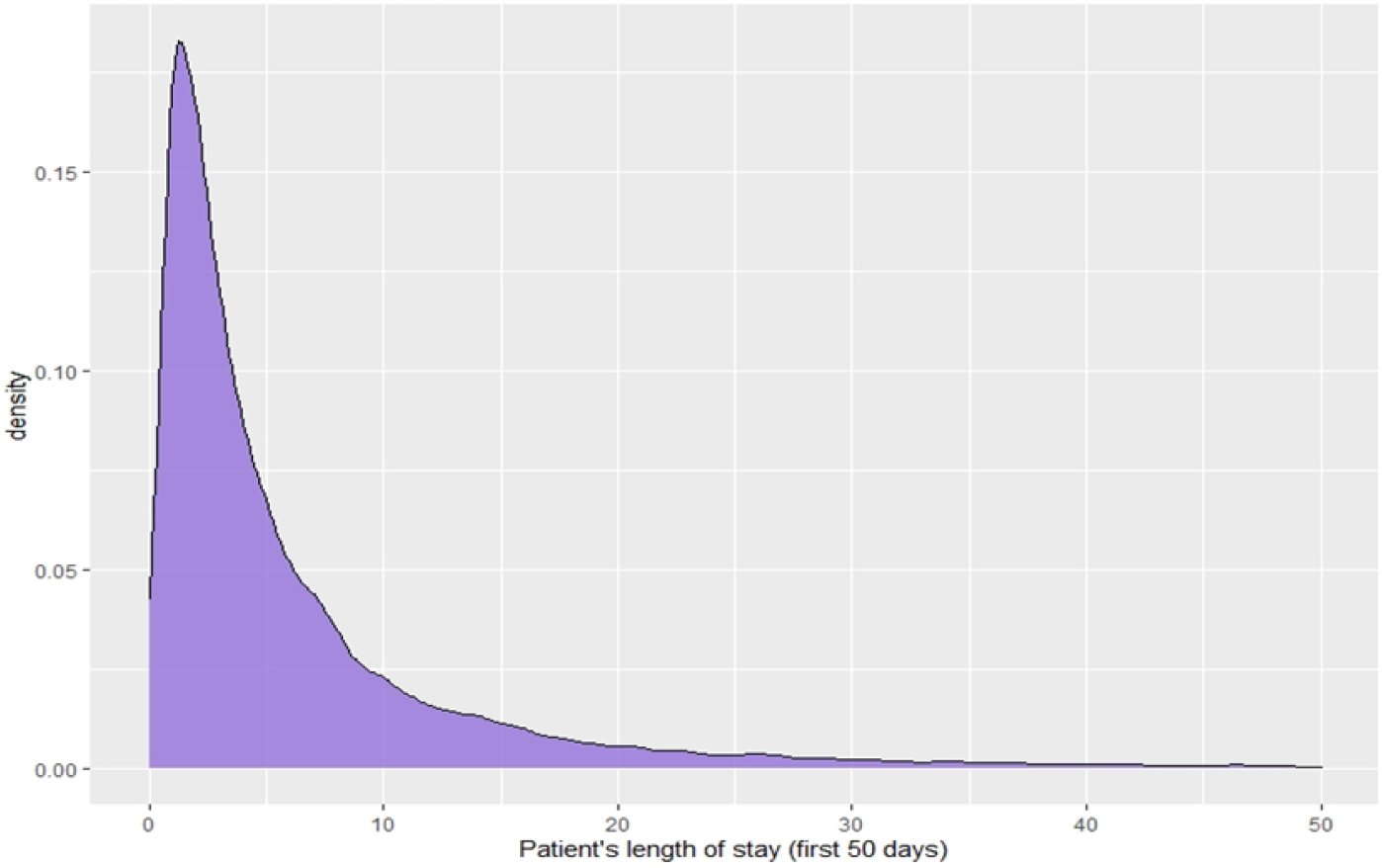
Length of stay for inpatient admission encounters (November 2018-2019) ***PLEASE USE COLOR FOR PRINTING FIGURE*** ***figure showing first 50 days from time of admission

There were a total of 43 adult inpatient clinical units, located in hospital 1 (n=18 units), hospital 2 (n=19 units), and hospital 3 (n=6 units). Hospital 1 accounted for 43.8% (n=16,438) of the admissions, while hospital 2 and hospital 3 constituted 37.4% (n=14,012) and 18.8% (7,062) of the admissions respectively. The majority of admissions were to high-complexity units, including NHPPD 6 units (66.7%, n=25039) and NHPPD 5.5 (14.6%, n=5460).

### 3.2 Completion and timeliness

Overall, the form completion rate across the three hospitals in the local health district was 78.4%. Hospitals 1, 2, and 3 had completion rates of 70.9%, 79.2%, and 91.2% respectively. As seen in Figure 2, the majority of forms are completed (78%, n=22,953) within the first 24 hours of admission. Other forms are completed between 24-72 hours (13%, n=3,910) or after 72 hours (9%, n=2,558) from the time of admission.

**Figure 2:**
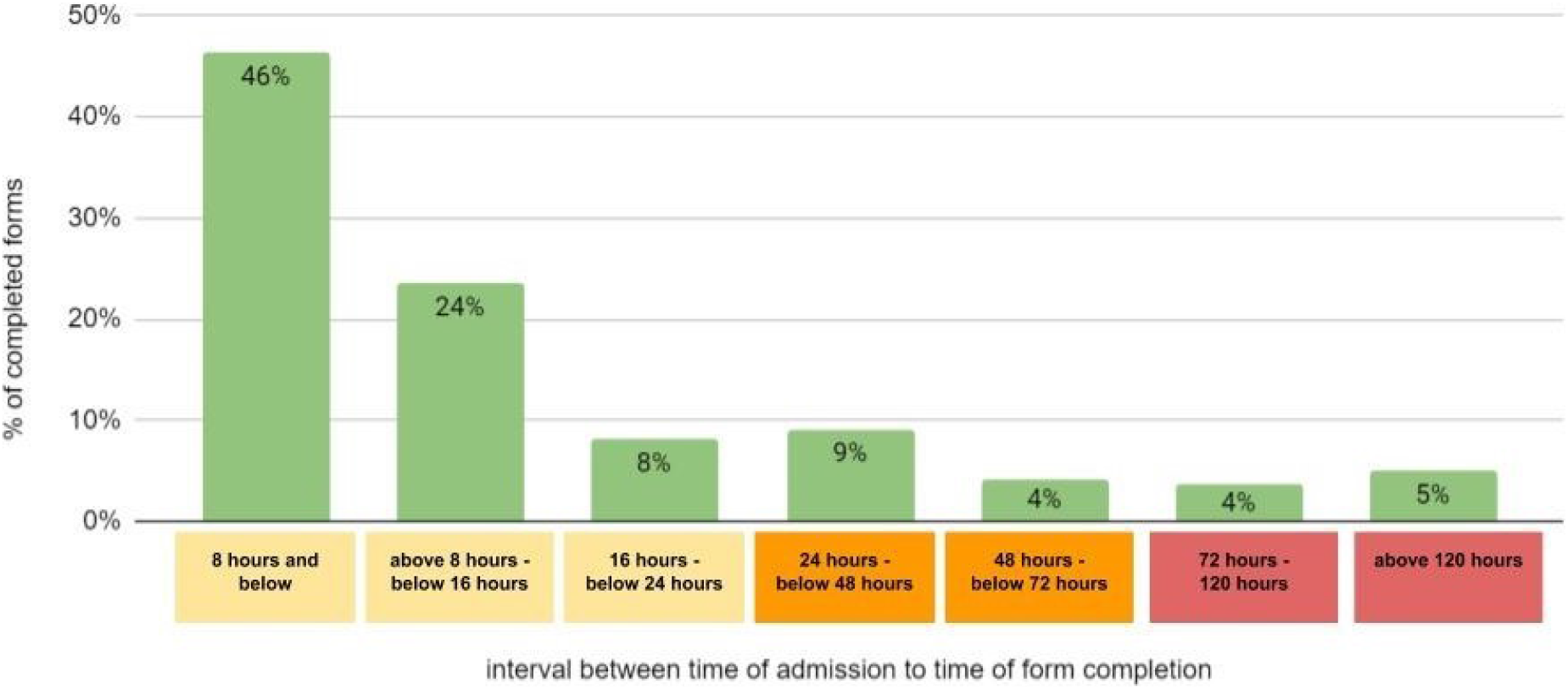
Timeliness of form completion ***PLEASE USE COLOR FOR PRINTING FIGURE***

### 3.3 Factors related to completion

Form completion was significantly more likely in the evening (aOR=1.13, 95% CI 1.06 – 1.21, p<0.001) than that in the morning.

Form completion was significantly less likely when length of hospital stay was shorter than 3 days (aOR 0.41-0.79, p <0.001), compared with those who stayed for 3 days. As the length of stay in hospital increased, form completion was observed to be more likely. Inpatient admissions with a duration of 5, 6, and 7 days had a 40-50% higher chance of form completion (aOR 1.40, 1.50, 1.37 respectively, p <0.001). Form completion was markedly higher for admissions beyond one week, expanding two-fold for those admitted up to 4 weeks (aOR=2.27, 95% CI 1.96 – 2.63, p <0.001), and five-fold for admissions more than 4 weeks (aOR= 5.55, 95% CI 4.39 – 7.01, p <0.001).

Form completion was marginally associated with NHPPD on an admitting clinical unit. Form completion for NHPPD 5.5 (aOR=2.74, 95% CI 1.03 – 7.33, *p*=0.044) and Non-NHPPD (aOR=2.95, 95% CI 1.03 – 8.44, *p*=0.044) units was significantly more likely compared with NHPPD 6 units. On the other hand, it was significantly less likely in a critical care unit (aOR 0.13, 95% CI 0.05-0.32, p <0.001). Inpatient admission encounters in clinical areas categorised as high acuity (CCU) and NHPPD 5 were nearly 60-90% more likely to have a completed form. Those in NHPPD 5.5 and Non-NHPPD clinical areas were three times more likely to have a form completed (aOR 3.02 and 3.42 respectively).

Patient gender and age group were not shown to be linked with form completion.

## 4. Discussion

Completed nursing documentation of admission assessment in the eMR is important because it allows nursing staff to assess, document and communicate relevant information and plan patient care accordingly. It is very important that this is done in a timely manner because safe, efficient, high-quality healthcare delivery from clinicians requires up-to-date information and movement of clinical data across and between care settings and time^12^. In acute/inpatient care settings, a clear overview of the patient is essential for multidisciplinary teams. Important relationships and trends might be overlooked if nurses are unable to document and view patient admission assessment data^26^.

To our knowledge, this is the first study addressing electronic admission assessment form completion among nurses in Australia. The high rate of form completion suggests that the majority of nurses comply with this admission-related task. A majority of patients have an AAA form completed within the first 8 hours of admission. While there are presently no mandated (or agreed) acceptable completion timeframes, this finding aligns with the general expectation that admission documentation should be done at the beginning of the patient’s hospital journey. It has been shown that timely communication and adequate nursing assessment are keys in preventing harm, detecting early deterioration, communicating a patient’s initial problems and needs and promoting patient safety.^2,27^

Though the high completion rates paint a fairly optimistic picture about electronic form completion, significant delays and non-completion rates were found for some admission encounters. Patient assessment is the primary reference for care planning and the start of the patient’s journey, yet in our study, one in five patients did not have an AAA form completed. This poses questions in relation to the process of care planning for acute care patients. Of those who had a form completed, almost a quarter were done 24 hours after admission, with 9% being done 5 days or more after a patient is admitted. The benefit of completing the form in the middle, or towards the end of a patient’s stay is questionable. In addition, this could suggest that the form may have been completed for administrative reasons rather than for patient care. This is counter to the principal purpose of nursing assessment and highlights a need for further education on the importance of timely documentation of patient assessment.

The inpatient admissions in this study were mostly from medical and surgical units. Urgency of direct patient care is a priority in these settings, staff busyness and/or patient acuity may compete with nurses’ inclination to complete electronic documentation. In a Middle Eastern study of factors contributing to nursing task incompletion, 780 medical and surgical nurses were surveyed and only 48% stated they could complete all the required procedures during their shift^28^. It is possible that nurses complete admission documentation outside of their shifts, as found in a Korean study which showed that nurses often stayed back to complete electronic documentation^11^. The round-the-clock nature of nursing shift work makes transitions and handovers vital to the completion of tasks, including admission documentation. It remains the case that while the eMR system could facilitate timely documentation, it should also be explored as a contributor to delays in form completion. The use of eMRs has been acknowledged as causative of work disruption, which may delay documentation of patient encounters^27^.

Form completion was found to be associated with the time of admission encounter. Prior studies^12,29,30^ have reported that the completion of electronic nursing documentation varies according to nursing shift, however, data in these studies were limited to only day and night shifts. Our study was able to address finer nursing shift arrangements that closely reflects our clinical practice (i.e. morning, evening, and night). In this study, evenings were the busiest shift, with nearly 43% of all inpatient admissions. This is similar to the findings of a North American study which explored nurse workload through clinical data, which revealed that the greatest activity in both the intensive care units and medical-surgical units occurred in the evening shift.^31^ Admission encounters during this time were also more likely to have a form completed, compared to those from a morning shift. In most public hospitals in New South Wales, the morning and evening shifts overlap as the former ends between 3pm and 4pm and the latter starts between 1pm and 2pm. This overlap of clinical staffing may contribute to increasing the likelihood of form completion during evening shift, as tasks are shared or delegated to incoming nurses. This study found that form completion was the lowest when patients were admitted in during the night shifts. This is consistent with a previous report that during night shifts, nurses spend less time in documentation than in day shifts.^11^ This is also in line with the fact that patients rest at night and patient assessment should ideally be completed at the bedside, with the patient fully awake.

Form completion was less likely in critical care and high complexity clinical areas. This may be explained by the increased nursing workload in these units, particularly as they are situated in acute tertiary hospitals. In an ICU work audit study, there were an average 178 nursing activities per patient per day in an intensive care setting, with 84% of activities performed by a single nurse.^31^ It may be that high workload among nurses in these high acuity clinical areas is a contributor to lower documentation. Another reasonable hypothesis here may be an anecdotal view that ICU nurses could privilege direct verbal communication over documentation-based communication more than their medical/surgical unit-based colleagues. Further, the existence of a parallel ICU-specific eMR likely explains some of the strength of this association. However, the parallel eMR did not contain all descriptors canvassed by the Cerner AAA form/was concerned more with patient progress than AAA descriptors.

This study found that patient’s gender and age are not associated with form completion. This finding contrasts that of another Australian study, which identified patients’ age as a factor significantly associated with healthcare form completion rate and timely documentation.^13^ This study examined a goals of care form, a resuscitation planning tool completed by medical staff to document informed decisions of hospitalised patients. Unlike the form we studied, such form is suited to an age-specific patient population who may require end-of-life conversations or palliative care planning and this may explain the difference in findings between the two studies.

The strength of this study lies on robust findings generated by strong statistical power, inclusion of multiple clinical areas, and use of multilevel modelling analytic techniques which accounted for hierarchies in the data. Its focus on the documentation of a single nursing process provides better insight into clinical practice compared with other studies which refer to electronic nursing documentation as a whole, or focus on physician practices alone. The inpatient admission encounters and data for analysis in this study were derived from an existing eMR audit report for the AAA form, so the variables included for analysis were limited. Nursing staff education on eMR use and practice variations could have influenced form completion, however such information was not available and lay outside the scope of this study. Future studies linking eMR data with reported critical incidents or harm to address outcomes of non-completion of the AAA form are recommended. Other patient-related characteristics such as Aboriginality, language spoken, primary diagnosis, or discharge disposition may provide additional insights into the factors influencing electronic form completion and nursing admission assessment.

This study investigated the timeliness of completing an electronic AAA form in a metropolitan Australian health district, and identified factors associated with its completion. Overall, form completion was high and often performed within the first 24 hours of hospital admission. Despite this, some patients did not have a form completed or form completion was significantly delayed. The time of admission, length of stay, and nursing hours per patient day in a unit were factors found to influence the completion of an AAA. More emphasis needs to be placed on the importance of completing the AAA form in a timely manner, as this informs initial care planning for the patient’s hospital stay. Education among nursing staff, exploration of reasons for form completion delay, and standardisation of nursing admission guidelines and eMR use are recommended for future practice.

## Supporting information

Author statement

## Data Availability

NA

## Authors’ contributions

DRS, GF, and DTT designed the study, performed the statistical analysis, and interpreted findings. DRS collected the data and wrote R code for cleaning and analysis. AJ contributed to interpretation of findings and provided executive support. All authors reviewed the final manuscript.

## Acknowledgment

The authors would like to thank Ivanka Komusanac, Executive Director of SLHD Nursing and Midwifery for her executive support. We would also like to thank SLHD Health Informatics, SLHD ICT Services, Sydney Research, and UNSW Centre for Big Data in Health Research for their kind support and assistance in this study.

