## Supplementary material for "Completion of electronic nursing documentation of inpatient admission assessment: insights from Australian metropolitan hospitals": Author statement

**Danielle Ritz Shala:** Conceptualization, Methodology, Software, Formal analysis, Data Curation, Writing-Original Draft, Writing-Review and Editing, Project administration, Visualization **Aaron Jones:** Resources, Supervision, Writing-Review and Editing **Greg Fairbrother:** Conceptualization, Methodology, Formal analysis, Writing-Review and Editing, Supervision **Duong Thuy Tran:** Conceptualization, Methodology, Formal analysis, Writing-Review and Editing, Supervision
